# Microsoft Bing vs. Google Bard in Neurology: A Comparative Study of AI-Generated Patient Education Material

**DOI:** 10.1101/2023.08.25.23294641

**Authors:** Talha Nazir, Usman Ahmad, Madho Mal, Mushhood Ur Rehman, Reeda Saeed, Junaid S. Kalia

## Abstract

**Background:** Patient education is an essential component of healthcare, and artificial intelligence (AI) language models such as Google Bard and Microsoft Bing have the potential to improve information transmission and enhance patient care. However, it is crucial to evaluate the quality, accuracy, and understandability of the materials generated by these models before applying them in medical practice. This study aimed to assess and compare the quality of patient education materials produced by Google Bard and Microsoft Bing in response to questions related to neurological conditions.

**Methods:** A cross-sectional study design was used to evaluate and compare the ability of Google Bard and Microsoft Bing to generate patient education materials. The study included the top ten prevalent neurological diseases based on WHO prevalence data. Ten board-certified neurologists and four neurology residents evaluated the responses generated by the models on six quality metrics. The scores for each model were compiled and averaged across all measures, and the significance of any observed variations was assessed using an independent t-test.

**Results:** Google Bard performed better than Microsoft Bing in all six-quality metrics, with an overall mean score of 79% and 69%, respectively. Google Bard outperformed Microsoft Bing in all measures for eight questions, while Microsoft Bing performed marginally better in terms of objectivity and clarity for the epilepsy query.

**Conclusion:** This study showed that Google Bard performs better than Microsoft Bing in generating patient education materials for neurological diseases. However, healthcare professionals should take into account both AI models’ advantages and disadvantages when providing support for health information requirements. Future studies can help determine the underlying causes of these variations and guide cooperative initiatives to create more user-focused AI-generated patient education materials. Finally, researchers should consider the perception of patients regarding AI-generated patient education material and its impact on implementing these solutions in healthcare settings.

## Introduction

The rapid advancements in artificial intelligence (AI) and natural language processing (NLP) have made it possible to create advanced language models that can produce content that resembles that of a human, like Google’s Bard and Microsoft’s Bing. These models are getting more and more complex, with potential uses in a range of industries, including healthcare (1). Patient education, which improves people’s understanding of their diseases, treatment plans, and health-related decisions, is a crucial component of healthcare. Healthcare practitioners can optimize information transmission and enhance patient care by using AI-driven language models to create patient education materials (2). However, it is crucial to take into account the quality, accuracy, and understandability of the designed materials (3) when applying AI-derived patient education tools in medical practice (4).

Microsoft Bing (5) and Google Bard (6) are two large language models that have access to real-time data and are also integrated or planned to be integrated in their respective search engines. This will ultimately change the behavior of users to search through these chatbots instead of traditional search. From the perspective of health education, patients may start using these chatbots to explain their symptoms or find information on specific diseases. In addition, clinicians should be aware of the limitations and use cases of these large language models to help patients find the best possible educational resources (7).

The majority of studies have focused on using ChatGpt in various areas of healthcare, such as medical education (4), clinical decision support systems (8), and board exam preparation (9). A comprehensive evaluation of Google Bard and Microsoft Bing’s capabilities for producing patient education content in the field of neurology is still lacking. By systematically assessing and contrasting the quality of patient education materials produced by Google Bard and Microsoft Bing, we hope to close this gap in our study. The findings of this study also provide a strong basis for the creation and continuous enhancement of AI-generated patient education materials in neurology and other branches of medicine.

## Objectives

1. Evaluate the quality of patient education materials generated by Google Bard and Microsoft Bing in response to questions related to neurological conditions.

2. Compare the performance of both AI models across key quality metrics, including accuracy, safety, comprehensiveness, objectivity, clarity, and compassion.

## Methodology

We have selected a cross-sectional study design for the purpose of evaluating and comparing Google Bard and Microsoft Bing’s ability to generate patient educational material.

For the purpose of this study, we have selected the top ten prevalent neurological diseases based on WHO prevalence data. The list of neurological conditions includes alzheimer’s disease, epilepsy, parkinson’s disease, multiple sclerosis, stroke, migraine, headache, motor neuron disease, spinal cord injury, and sciatica. The selection was made with the intention of covering a wide spectrum of disorders that patients regularly deal with. We developed a patient-centered prompt question for each neurological disease. The prompts were designed to be comprehensive, objective, and specific. We have then generated patient education material based on these prompts by using Google Bard and Microsoft Bing. For each prompt, we have created a new chat to avoid any impact from previous prompts. The created content was gathered and arranged for assessment.

Ten board-certified neurologists and four neurology residents were asked to evaluate these responses on a five-point Likert scale, with 1 being the lowest and 5 being the highest. Their varied backgrounds helped create a more thorough and unbiased evaluation. Six quality metrics were made available to the assessors: accuracy, safety, comprehensiveness, objectivity, clarity, and compassion. The name of the AI model connected to each response was hidden from the participants in order to reduce bias.

The data was analyzed using IBM SPSS Statistics software. To determine an overall score for each language model, the scores for each model have been compiled and averaged across all measures. The significance of any observed variations across the models was then assessed using an independent t-test. We performed additional analyses, looking at the results for each condition and contrasting the performance of the AI models for specific queries, to learn more about the efficiency of both AI models. Using this method, we were able to pinpoint particular situations in which one model might produce high-quality patient education materials more effectively than another.

### Results

We evaluated the performance of Google Bard and Microsoft Bing in generating patient education For nine neurological diseases, we compared how well Google Bard and Microsoft Bing produced patient education items. Due to Google Bard’s inability to produce an answer, the migraine query was removed from our analysis, leaving nine prompts for comparison.

### Overall Performance

The mean scores for each model were 79% for Google Bard and 69% for Microsoft Bing. While comparing two models, Google Bard outperformed Microsoft Bing in all six-quality metrics, with overall scores of 81% versus 71% for accuracy, 79% versus 69% for safety, 79% versus 67% for comprehensiveness, 77% versus 69% for objectivity, 79% versus 70% for clarity, and 75% versus 67% for compassion. The independent t-test showed a significant difference between the two models for all metrics (p < 0.05). Detailed metric-wise comparison results are presented in Figure 1 and Supplementary Material Figure 2.

**Figure 1:**
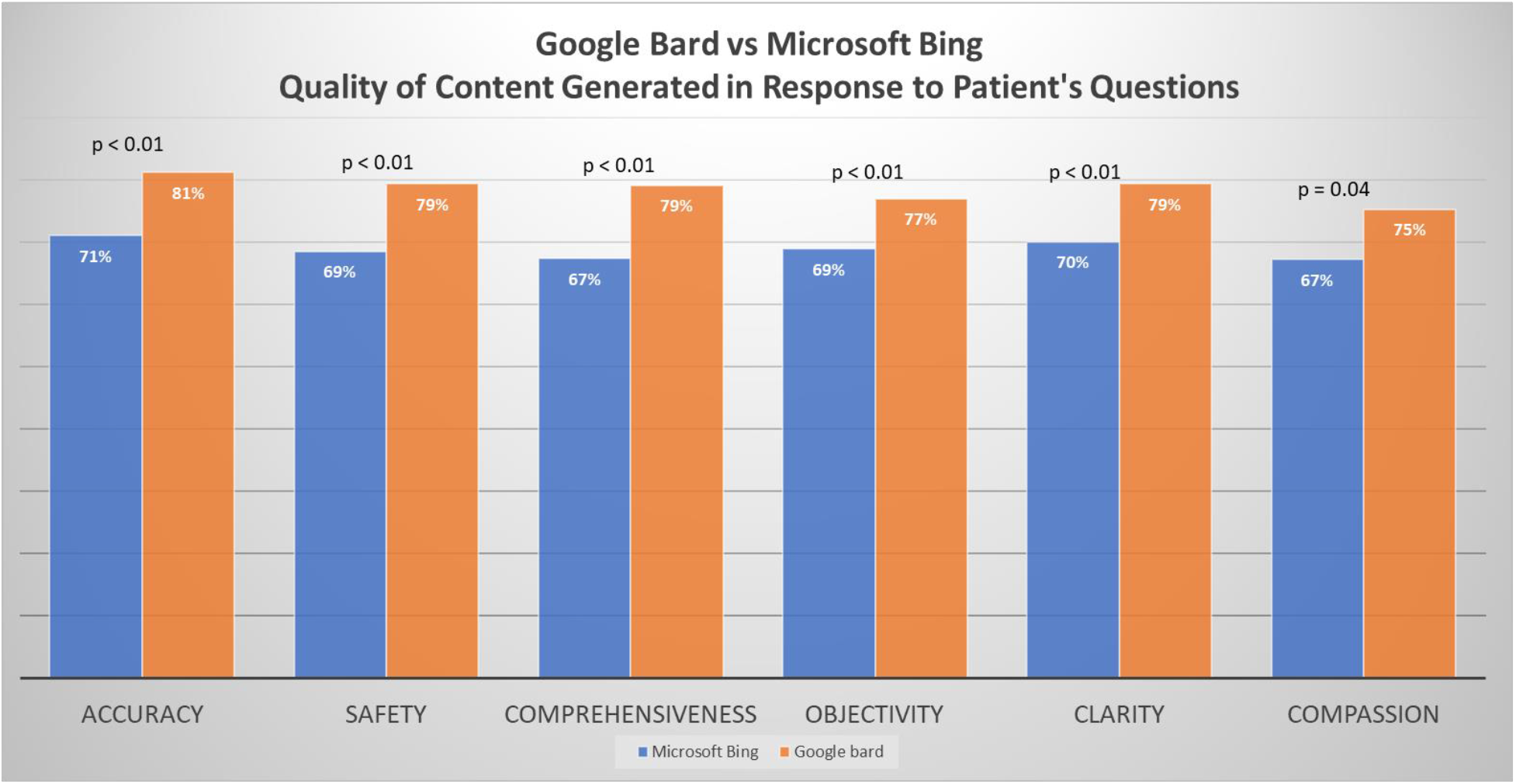
Comparison of Microsoft Bing and Google Bard.

**Figure 2:**
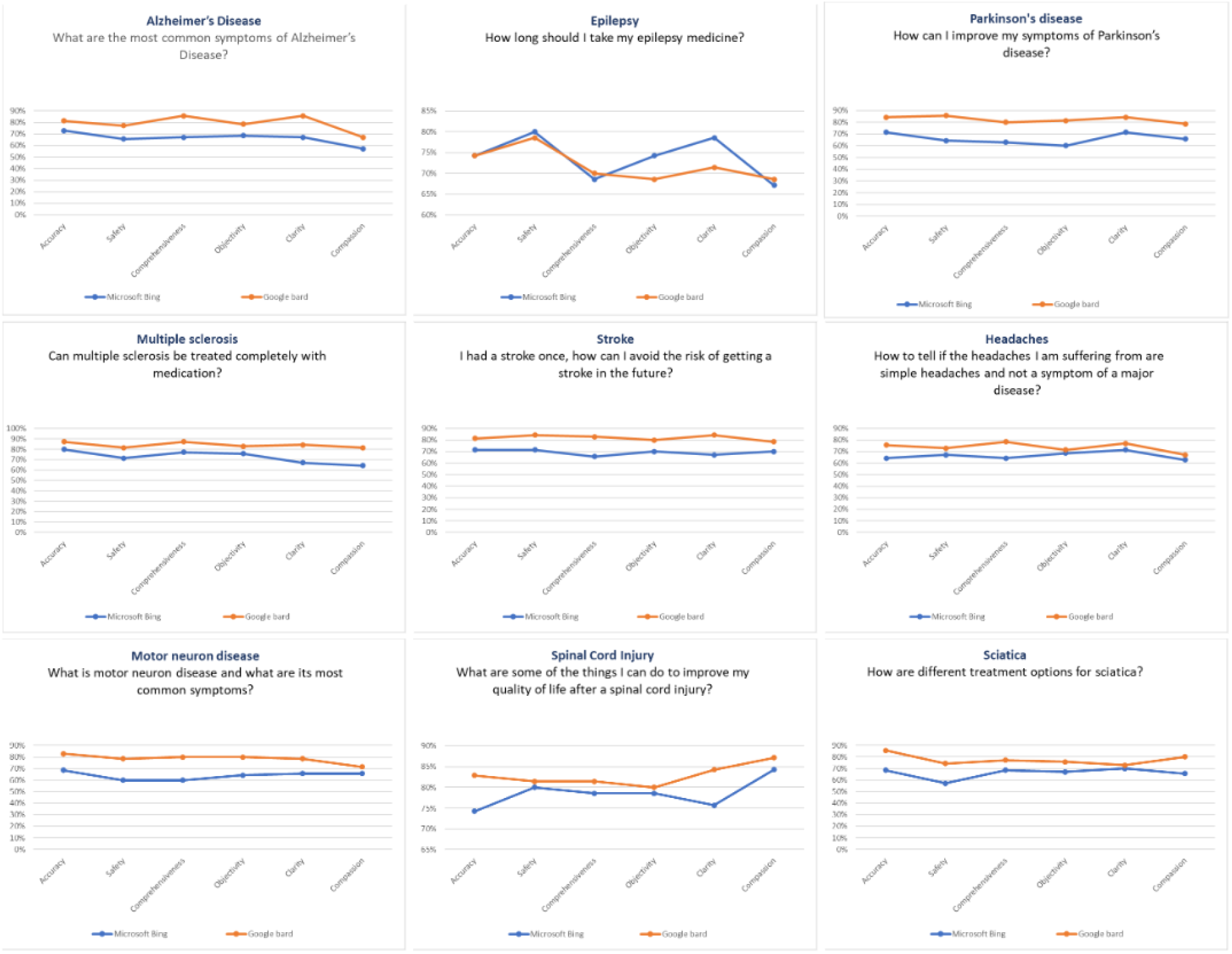
Performance of Microsoft Bing and Google Bard based on individual question.

### Performance by Individual Neurology Questions

By looking at the results of specific questions, we performed a more thorough analysis of the effectiveness of both AI models in producing patient education materials. For eight neurological illnesses, including sciatica, stroke, motor neuron disease, spinal cord injury, multiple sclerosis, Parkinson’s disease, Alzheimer’s disease, and headache, Google Bard exceeded Microsoft Bing across all six quality categories (see Figure 2). For the epilepsy query, Microsoft Bing performed marginally better in terms of safety, clarity, and objectivity. This implies that while Google Bard is generally better at producing high-quality patient education resources, there are particular situations where Microsoft Bing’s content might also offer helpful information.

These findings emphasize the significance of taking into account each AI model’s limitations and strengths when utilizing them to produce patient education materials because their efficacy may differ based on the particular context and inquiry.

## Discussion

Our study showed that Google Bard performs better than Microsoft Bing in six quality categories, with an overall mean score of 79% and 69%, respectively. In addition, Google Bard outperformed in all six measures for eight questions, according to the analysis of each question separately. For the epilepsy query, Microsoft Bing performed marginally better in terms of objectivity and clarity. When using AI-generated content for patient education, healthcare providers should account for these variations in order to maintain the safety and quality of the content. Our study is distinctive in that it compares Google Bard’s and Microsoft Bing’s performance, offering useful information for healthcare professionals thinking about using AI-generated material for patient education.

The already published literature is mostly focused on the use of ChatGpt (Microsoft Bing) in generative patient education material in different specialties of medicine, such as gastroenterology (10), cardiology (11), bariatric surgery (12, 13), otolaryngology (14), ophthalmology (15), and prostate cancer (16). In our study, we have not only evaluated the Microsoft Bing and Google Bard models individually but also compared them in their ability to produce quality patient education content.

The limitations of this study include the inclusion of a relatively small number of neurological disorders and questions evaluated. In addition, the sample size was also small. Future studies can minimize these limitations by looking at how well AI models perform in producing patient education materials across a wider range of medical specialties and with more prompts and conditions. Researchers should also consider the factors (quality of training data, real-time web access) that distinguish one AI model from another in specific health information fields. By taking these elements into account, search engine providers and AI developers could collaborate to produce specialized solutions to cater to individual patients’ needs.

Furthermore, researchers may also consider the perception of patients regarding AI-generated patient education material and its impact on implementing these solutions in healthcare settings. The performance and protocol of these technologies can be improved by understanding the needs and opinions of patients, which will ultimately improve the efficacy of patient education.

Finally, as AI technology develops, it is critical to make sure that the patient education materials produced by these models are relevant, culturally appropriate, and morally sound. This can entail putting in place systems for ongoing assessment and feedback, as well as including patients and subject-matter experts in the evaluation and improvement process.

## Conclusion

Our research advances knowledge on the quality of patient education materials produced by Google Bard and Microsoft Bing, particularly in the context of neurological diseases. In the majority of quality metrics, Google Bard performs better than Microsoft Bing; however, healthcare professionals should take into account both AI models’ advantages and disadvantages when providing support for certain health information requirements. Additional studies can help determine the underlying causes of these variations and guide cooperative initiatives to create more user-focused AI-generated patient education materials.

## Data Availability

All data produced in the present study are available upon reasonable request to the authors.

